# PATIENT PATHWAYS TO UGANDA’S FIRST SPECIALISED EARLY INTERVENTION IN PSYCHOSIS SERVICE AND RELATION TO THEIR CLINICAL OUTCOMES

**DOI:** 10.64898/2026.04.30.26352152

**Authors:** Emmanuel Kiiza Mwesiga, Wilber Ssembajjwe, Rossette Immy Ndigamanya, Sophia Balinga, Blessed Tabitha Aujo, Mary Ampaire, Andrea Kaggwa Kaddu, Andrew Sentoogo Ssemata, Allan Kalungi, Ronald Kiguba, Johesephat Byamugisha, Mark Kaddumukasa, Martha Sajatovic, Noeline Nakasujja

## Abstract

**Background:** Early Intervention for Psychosis Services (EIPS) enhance outcomes for individuals experiencing their first episode of psychosis (FEP). However, in low-resource settings, there is limited knowledge about i) the pathways patients take to access EIPS, ii) the proportion and factors associated with acceptance of referral to EIPS, and iii) if different pathways to EIPS services affect clinical outcomes. Uganda’s first EIPS, the Specialised Treatment Early in Psychosis Service at Makerere University Hospital (STEP_MaKH), presents a unique opportunity to explore these important questions.

**Aims:** We aimed to examine the pathways to EIPS, the factors associated with referral to specialised psychosis care and the impact of initial treatment-seeking behaviour on long-term symptom remission and quality of life.

**Methods:** We conducted a multiple-method study. Pathways to care were assessed retrospectively using the WHO Encounter Form among adults with FEP eligible for referral to STEP_MaKH. Among those who completed referral and enrolled in STEP_MaKH. Symptom severity and quality of life were followed prospectively for 12 months. Modified Poisson regression identified predictors of referral completion. Kaplan–Meier methods and Cox proportional hazards models examined time to symptom remission and time to achieving a good quality of life.

**Results:** Of the 187 adults with first-episode psychosis eligible for referral to STEP_MaKH, Native/religious healers (n = 86) were the predominant first point of contact. Only 56 (29.9%) accepted referral to STEP_MaKH. Participants referred from Mulago National Referral Hospital more likely to enrol than those referred from Butabika (RR = 4.7; 95% CI: 2.90–7.87). Longer delays from first treatment contact were associated with reduced likelihood of reaching STEP_MaKH (RR = 0.99 per month; p = 0.041). After enrolment, symptoms improved rapidly with 60% achieving PANSS remission by Month 1, and fewer than 10% remained non-remitted by Months 2–3. In adjusted Cox models, participants initially seen by mental health workers achieved remission more quickly than those initially seen by non-medical personnel (HR = 1.48; 95% CI: 1.05–2.10). Older age was associated with slower remission (HR = 0.94; p = 0.023). Quality of life improved over the follow-up period, with earlier attainment of good quality of life among those initially managed by mental health workers.

**Conclusions:** Pathways to care for FEP in Uganda are complex and culturally mediated, with substantial attrition before specialised early psychosis care is reached. Referral completion is strongly shaped by referral site and by delays in the care pathway. Once in specialised care, clinical outcomes improve rapidly, and initial contact with mental health workers is associated with faster symptom remission and earlier gains in quality of life. Strengthening referral systems, reducing pathway delays, and developing collaborative detection-and-referral links with community and frontline providers are key priorities for optimising early psychosis outcomes in low-resource settings.

## Introduction and Background

First-episode psychosis (FEP) refers to the initial presentation of psychotic symptoms, typically characterised by delusions, hallucinations, disorganised thinking, and behavioural disturbances occurring for the first time in an individual’s life (1–3). This phase represents a critical and potentially reversible period in the trajectory of psychotic disorders, during which timely intervention is associated with improved symptomatic, functional, and quality-of-life outcomes (2, 4). Unlike chronic schizophrenia, FEP is marked by greater neurobiological plasticity, lower cumulative treatment exposure, and a higher probability of remission, making early detection and intervention particularly consequential (5, 6). Delays during this stage, often conceptualised as duration of untreated psychosis (DUP), are strongly associated with poorer long-term prognosis (6, 7).

It is recommended that patients experiencing their first-episode psychosis (FEP) receive initial care in an early intervention for psychosis service (EIPS) (6, 8, 9).Evidence-based interventions, including low-dose second-generation antipsychotics, psychotherapy, and supported education and employment, improves outcomes in FEP Treatment of FEP in EIPS is associated with better outcomes, including quality of life, duration of hospitalization and symptom remission in high income countries (10–14). The effectiveness of EIPS has shifted policy frameworks globally, positioning early intervention as a cornerstone of modern psychosis care.

The pathways patients take to reach EIPS vary widely and are influenced by health system structures, cultural beliefs, and healthcare accessibility (15, 16). In high-income settings, studies have mapped these pathways, demonstrating that early access to specialised care reduces the duration of untreated psychosis and improves clinical and functional outcomes (8, 17).

Despite strong evidence from high-income settings, the implementation and evaluation of EIPS in low- and middle-income countries (LMICs), particularly in sub-Saharan Africa, remain limited. The few available studies in many low-resource settings, particularly in sub-Saharan Africa, document that pathways to care are often non-linear, and shaped by cultural explanatory models, stigma, and constrained mental health infrastructure. Patients often seek for help from multiple providers, including traditional healers and general medical services, before reaching psychiatric care (18–23). Delays in accessing appropriate treatment contribute to poorer long-term outcomes, underscoring the need to understand referral dynamics in these settings (24–26). Empirical data on how patients reach EIPS and whether these pathways influence clinical outcomes in Sub-Saharan Africa are scarce.

Understanding patient trajectories, defined as the sequence and transitions between care providers from symptom onset to specialised treatment, is essential for evaluating real-world access to and effectiveness of EIPS. Care pathways are not merely descriptive phenomena; they influence delays, treatment exposure, family responses, and ultimately clinical outcomes. Mapping trajectories allows identification of system bottlenecks, attrition points, and inequities in referral mechanisms (15, 17). In emerging EIPS models within LMICs, analysing trajectories is particularly important to distinguish between service efficacy and access limitations.

Given the scarcity of data on EIPS is Uganda and SSA more broadly, we conducted a retrospective descriptive analysis of clinical care pathways taken by 187 patients with first episode psychosis over a 12-month time-frame within a health care system in Uganda. We describe the referral pathways to Uganda’s first EIPS, the Specialised Treatment Early in Psychosis Service at Makerere University Hospital (STEP_MaKH), and examine their impact on quality of life and symptom remission. By mapping patient trajectories, we aim to1) examine the pathways to EIPS, 2) the factors associated with referral to specialised psychosis care and 3) the impact of initial treatment-seeking behaviour on long-term symptom remission and quality of life. Understanding these dynamics is critical for designing interventions that reduce delays and improve engagement with EIPS.

## Methods

### Study design

The study employed a multiple-method design, with retrospective assessments to determine pathways to care and prospective measures to examine relationships between different pathways with quality of life and symptom remission among 187 patients with FEP during their first year of treatment in a Ugandan health system.

### Study setting

Mental health services in Uganda are delivered through a tiered system comprising primary care facilities, general hospitals, and specialised psychiatric hospitals. Public specialist psychiatric care is concentrated in a limited number of referral institutions, including Butabika National Referral Mental Hospital, while integration of mental health into general medical services remains variable. Access to care is influenced by workforce shortages, stigma, geographical barriers, and pluralistic health-seeking practices, with many individuals initially consulting traditional or religious healers prior to engaging with formal psychiatric services (27, 28). Mulago National Referral Hospital and Butabika National Referral Mental Hospital serve as major entry points for patients with severe mental illness, including first-episode psychosis.

Participants were recruited from Butabika National Referral Mental Hospital, Mulago National Referral Hospital, and affiliated clinical services. Individuals presenting with symptoms consistent with first-episode psychosis were screened for eligibility using diagnostic criteria and inclusion/exclusion parameters. Eligible participants underwent retrospective assessment of pathways to care. All eligible individuals were offered referral to STEP_MaKH. Participants who accepted referral and completed enrolment entered the prospective follow-up cohort and were assessed longitudinally over 12 months for symptom severity and quality-of-life outcomes.

STEP_MaKH is a pioneering mental health service in Uganda, established in 2024, that aims to promote the early identification, intervention, and ongoing care of individuals experiencing their first episode of psychosis (FEP) (29)(ref). It was established to address critical gaps in early psychosis care in Uganda. The clinic offers specialised assessment, diagnosis, and treatment. Care is delivered by a team that includes psychiatrists, clinical psychologists, and psychiatric nurses, each with a small patient caseload. The model emphasises family involvement, psychoeducation, psychopharmacology, and, where feasible, psychosocial interventions. The clinic primarily receives referrals from Butabika and Mulago National Referral Hospitals. STEP_MaKH is increasingly integrating measurement-based care tools, including components from EPINET’s Core Assessment Battery, to monitor patient progress, guide treatment decisions, and enhance quality improvement (30, 31).

### Participants

The study included first-episode psychosis patients operationalised as having a confirmed psychosis disorder according to the MINI International Neuropsychiatric Inventory (MINI)(32). The sample size of 187 participants included in the analysis, were adults aged 18-60, never having been treated with antipsychotic medication or on medication for less than six weeks’ duration and no substance use disorder, HIV/AIDS or syphilis.

### Variables and data sources

Pathways to care were identified using the World Health Organisation’s Encounter form to systematically gather information on the sources of care utilised by participants before approaching a mental health professional (33). Participants were then offered a referral to STEP_MaKH. Outcome measures were collected only among participants enrolled in STEP_MaKH and were administered as part of routine measurement-based care at baseline (enrolment) and at Months 1, 2, 3, 6, and 12. Participants who agreed to referral to STEP_MaKH received routine specialised care, including repeated measures to track outcomes from the EPINET core assessment battery (34, 35). The battery includes the six-item Positive and Negative Signs and Symptoms of Schizophrenia (PANSS-6) scale to assess symptom severity and the EPINET Quality of Life Tool to assess quality of life. These have been described elsewhere.

### Statistical analysis

Categorical variables such as sex, religion, district of residence, employment status, and type of first contact were summarized using frequencies and percentages. Continuous variables, including age, were summarized using means and standard deviations. To compare participants who received specialized psychiatric care (STEP_MaKH) with those who did not, Pearson’s Chi-square tests were used for categorical variables and independent-sample t-tests were used for continuous variables. Fisher’s exact tests were applied where expected cell counts were small. Statistical significance was set at p < 0.05.

A multivariate modified Poisson regression model with robust standard errors was fitted to identify factors independently associated with referral completion to the Specialised Treatment Early in Psychosis Service (STEP_MaKH). Independent variables included demographic characteristics, recruitment site, and type of first contact. Adjusted risk ratios (RRs) with 95% confidence intervals (CIs) were reported. Care-seeking trajectories were examined using the World Health Organisation (WHO) Encounter Form, which captures sequential points of contact across different providers. Pathways were categorised by type of first contact (traditional or religious healer, general hospital, police, social worker, or psychiatric service) and mapped to final presentation at Butabika National Referral Mental Hospital (BNRMH) or STEP_MaKH. The number and proportion of individuals following each unique pathway were calculated and summarised separately for the overall sample, those enrolled in EIPS (STEP_MaKH), and those who remained at BNRMH. Visual flow diagrams were used to illustrate common referral patterns and identify variations in access to psychiatric care.

Longitudinal changes in clinical outcomes were analysed for participants followed at STEP_MaKH. The outcomes were plotted across four time points (t₀ to t₃) and stratified by initial point of contact, traditional/religious healers versus medical and mental health providers. These visualisations were used to describe differences in trajectories of symptom remission and quality of life over time. Finally, survival analyses were performed to assess the time to PANSS remission and achieving satisfactory quality of life (defined as QOL ≥7 - defined by principal component analysis). Kaplan–Meier survival curves were generated to estimate the cumulative probability of achieving satisfaction over time. Covariate-adjusted hazard ratios (HRs) were obtained using Cox proportional hazards models, with age, sex, BMI, and PANSS included as predictors. Model diagnostics were assessed using log-rank tests and the proportional hazards assumption. All statistical analyses were conducted using Stata version 18 (StataCorp LLC, College Station, TX, USA) and SAS 9.4M6

### Ethics and Dissemination

Ethical clearance was obtained from the School of Medicine Research and Ethics Committee (SOMREC) (Mak-SOMREC-2024-869) and institutional approval from Butabika National Referral Mental Hospital and Makerere University Hospital. All participants provided informed consent prior to participation.

## Results

A total of 187 participants were recruited for the study, with 85.5% (160/187) recruited from Butabika National Referral Mental Hospital. The median age of the participants was 28.9 years, and most (60%) were males. Most participants were from Wakiso (46%) and Kampala (31%), with smaller proportions from Mukono and other districts. The majority were Christian (81%), followed by Muslims (17%), and a small number from other religions. In terms of education, most had completed primary (35%) or secondary (39%) education, while smaller numbers had tertiary (22%) or no formal education (5%)

### Pathways to care

This figure illustrates the multiple, often cyclical care-seeking paths individuals take before reaching formal psychiatric services. Native/religious healers (n = 86) are the predominant first point of contact. Notably, a large proportion of participants used general psychiatric services as their initial point of care for the first episode of psychosis (n = 61). Other initial points of contact were established with social workers (n = 1), police (n = 7), and general hospitals (n = 30). Following that initial contact, the patients continued to seek care from various service providers, including traditional or religious healers. For example, of the 86 individuals who first received care from native/religious healers, 27 continued care with another native/traditional healer, 11 went to a general hospital, 38 to a psychiatric hospital, and one accepted to continue care with STEP_MaKH. Other pathways to specialised care are shown in the figure below Referral to STEP_MaKH: The pathways to STEP-MaKH were not direct for some individuals. Although more than half of the participants reached STEP_MaKH via a single referral, some required two or three referrals, as shown in the figure below. For example, the participant who first contacted the police required three referrals to reach STEP_MaKH. The one participant whose initial contact was a social worker never made it to the specialised clinic.

**Figure 1:**
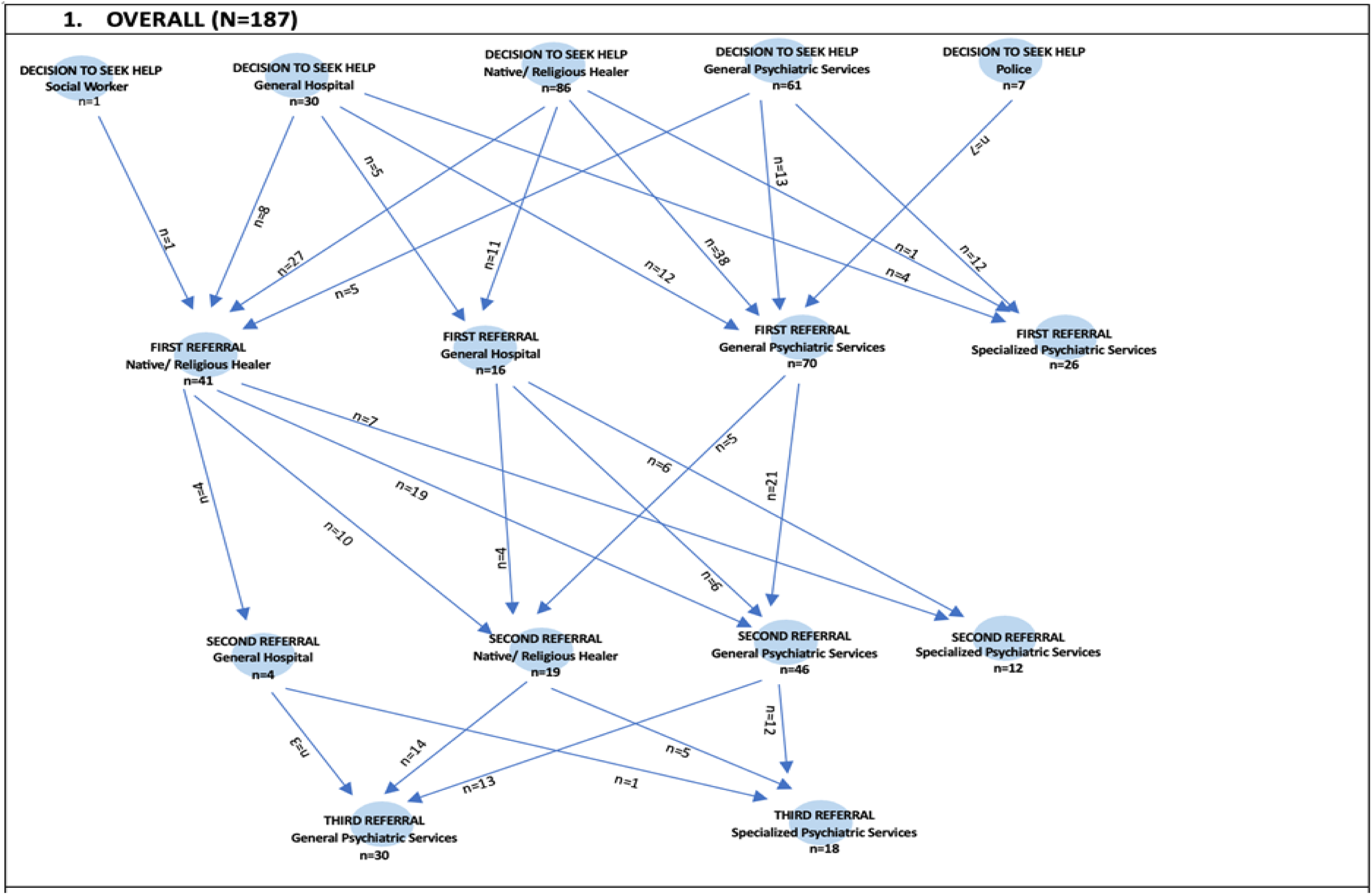
Pathways to care for the whole sample.

**Figure 2:**
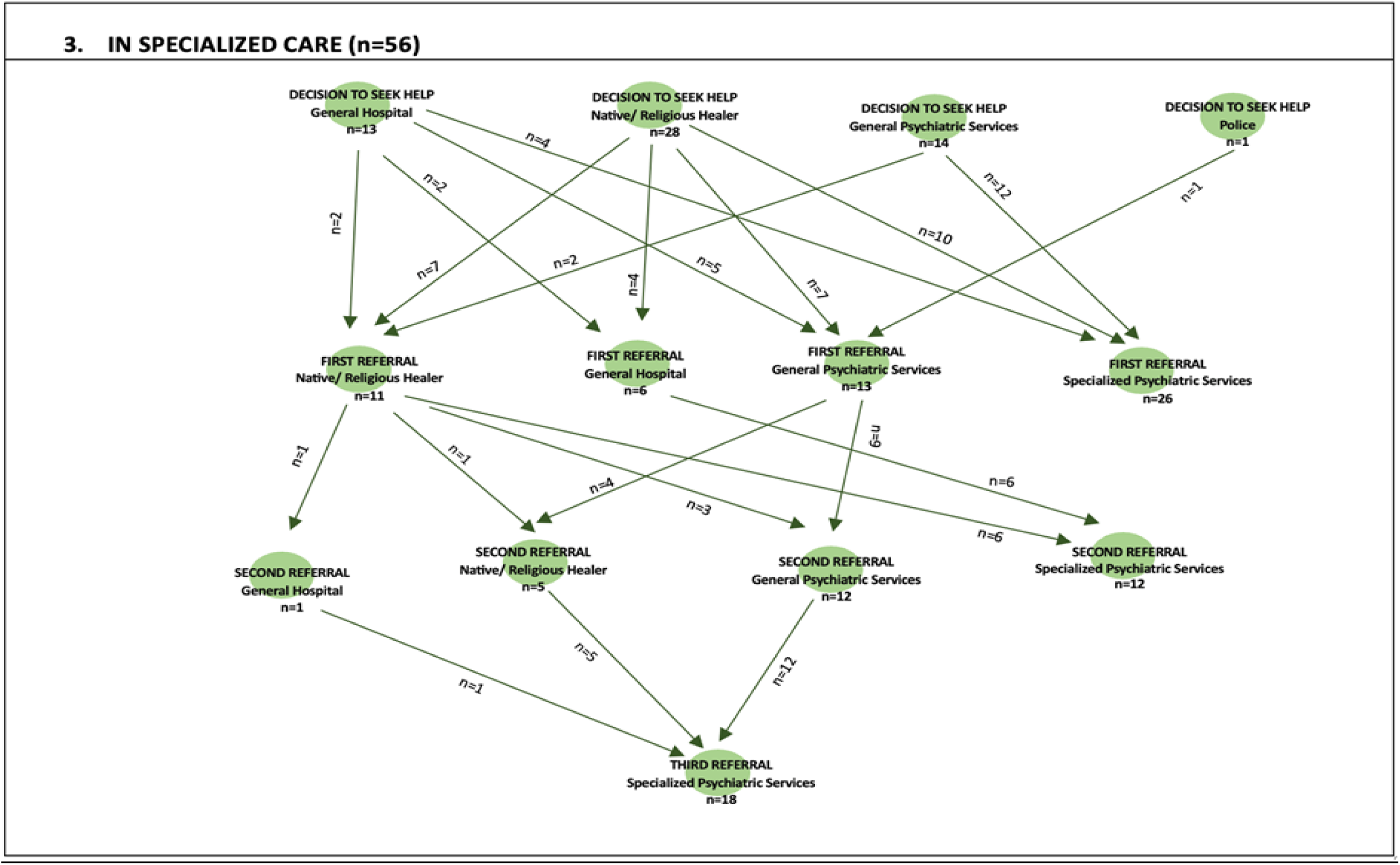
Pathways to care to STEP_MaKH.

Only 56 of the 187 participants agreed to a referral to the STEP_MaKH. Participants who accepted referral to STEP_MaKH were slightly younger, with a mean age of 27.2 years (SD = 7.9), compared with 30.5 years (SD = 9.4); this difference was statistically significant (p = 0.023). There were also statistical differences among those who reached STEP_MaKH by referral source and educational level. Other comparisons are shown in the table below. Among those who received specialised care, most were from Butabika National Referral Hospital (32/56; 57.1%). However, a higher proportion of participants (16/18, 88.9%) accepted referral to STEP_MaKH from Mulago NRH compared to only 32/160 (20.0%) from Butabika Hospital. Further distribution of participants who accepted referral compared to those who didn’t across various sociodemographic characteristics are shown below.

**Table 1:**
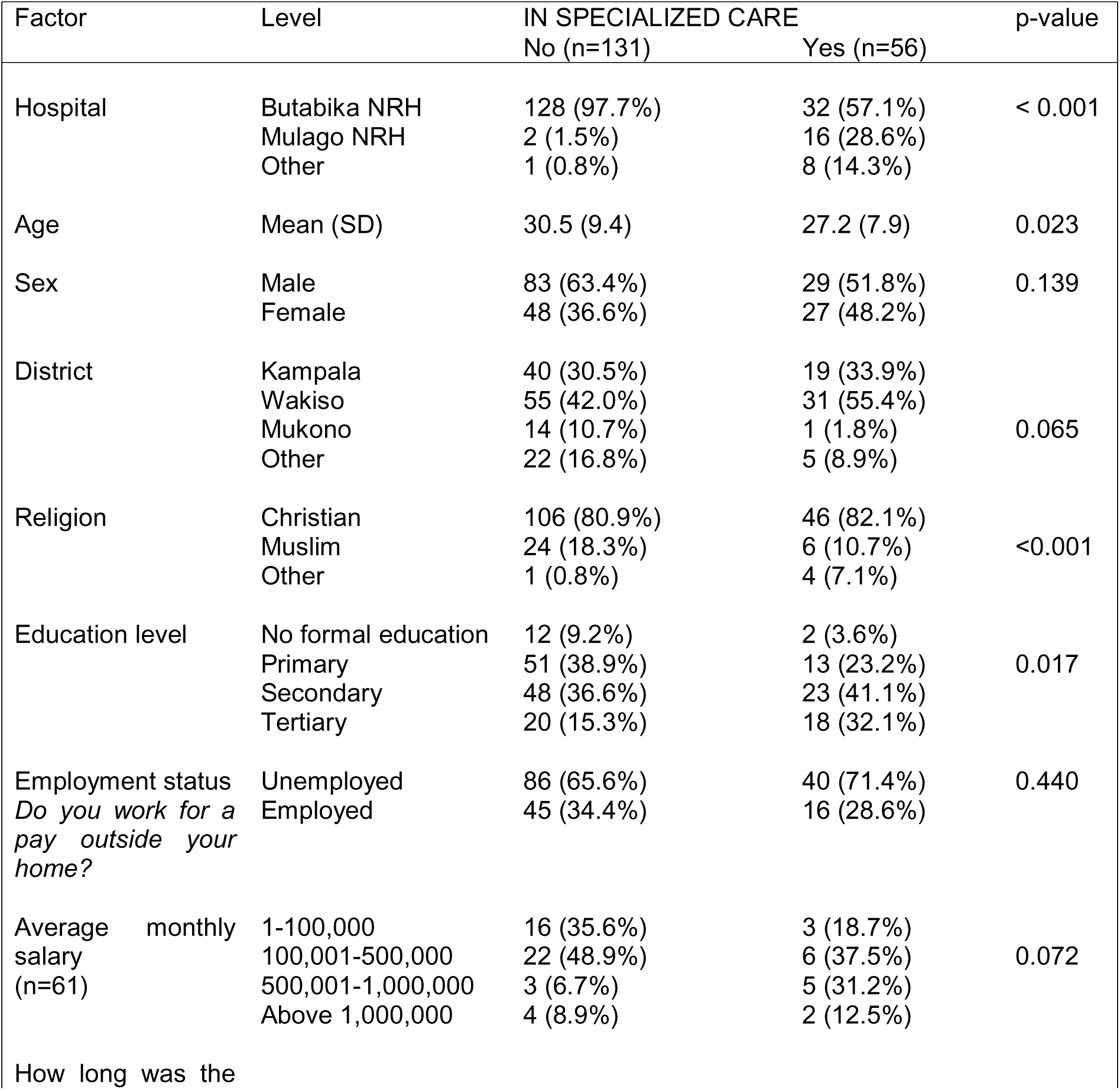
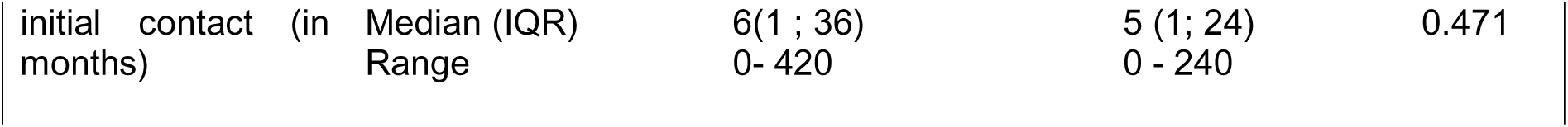
Demographic Characteristics.

In the multivariate modified Poisson regression model, the predictors of accepting referral to STEP_MaKH included the referral site, with participants referred from Mulago National Hospital four times more likely to make it to STEP_MaKH than those from Butabika National Referral Mental Hospital (RR 4.7 (2.90;7.87, P<0.001). Furthermore, the person initially contacted and the time since contact were also associated with a reduced risk of presenting to STEP_MaKH, as shown in the table below.

**Table 2:**
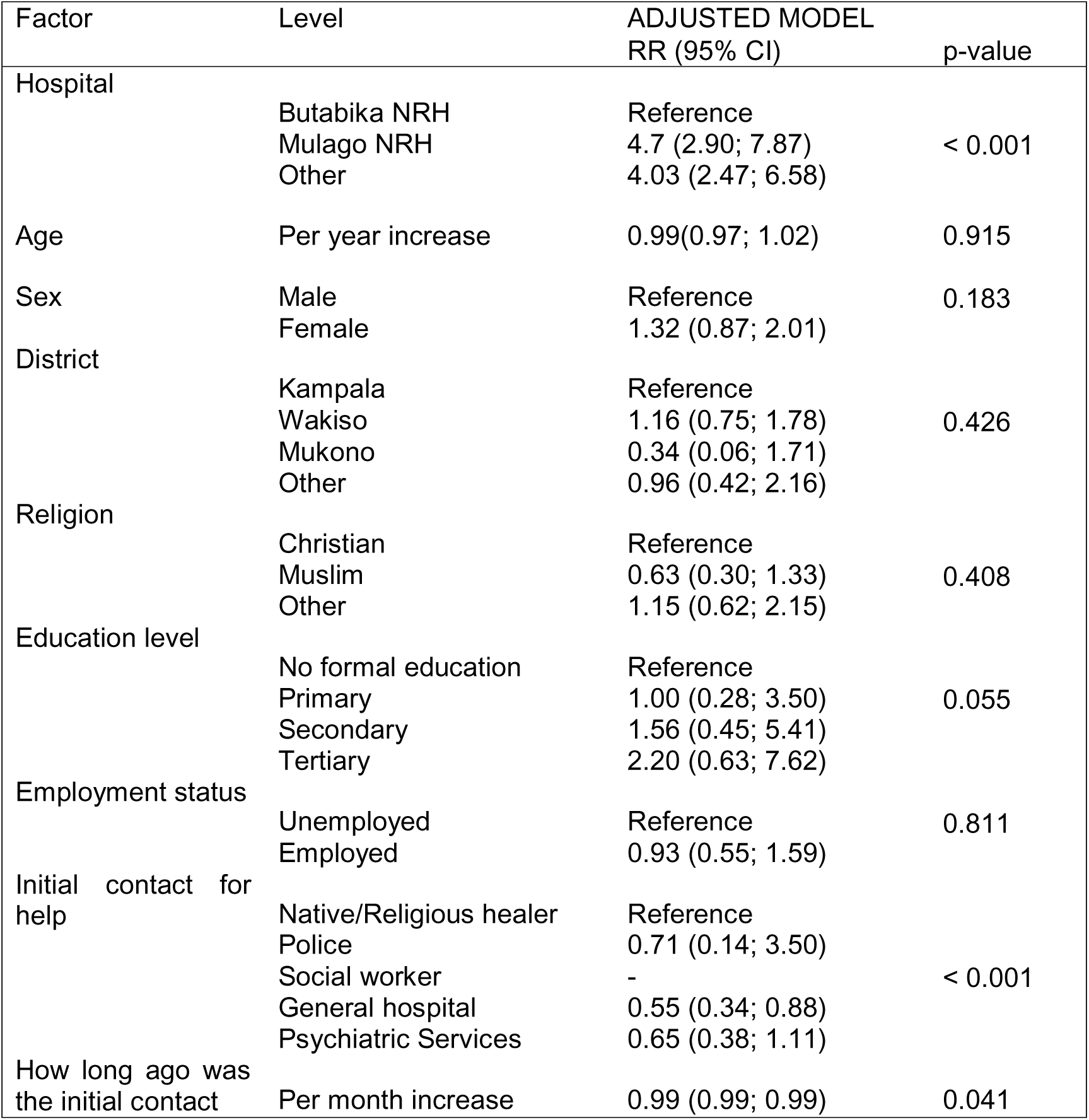
Results for fitting a multivariate modified Poisson model.

### Initial point of care and patient outcomes

**Symptom remission** (PANSS-6 score <14): At baseline, mean PANSS scores were highest among participants initially seen by mental health workers, followed by those seen by non-medical personnel and general medical doctors. Between baseline and Month 2, all groups showed a substantial reduction in mean PANSS scores, with values declining to approximately 7.4–7.8 across groups. Further reductions were observed at Month 3 and Month 6. At Month 6, mean PANSS scores were lowest across the follow-up period, at approximately 6.3 for participants initially seen by mental health workers, 6.5 for those seen by general medical doctors, and 6.7 for those seen by non-medical personnel. By Month 12, mean PANSS scores among participants initially seen by mental health workers continued to decline slightly, while scores among those initially seen by general medical doctors remained relatively stable and scores among those initially seen by non-medical personnel increased modestly

**Figure 3:**
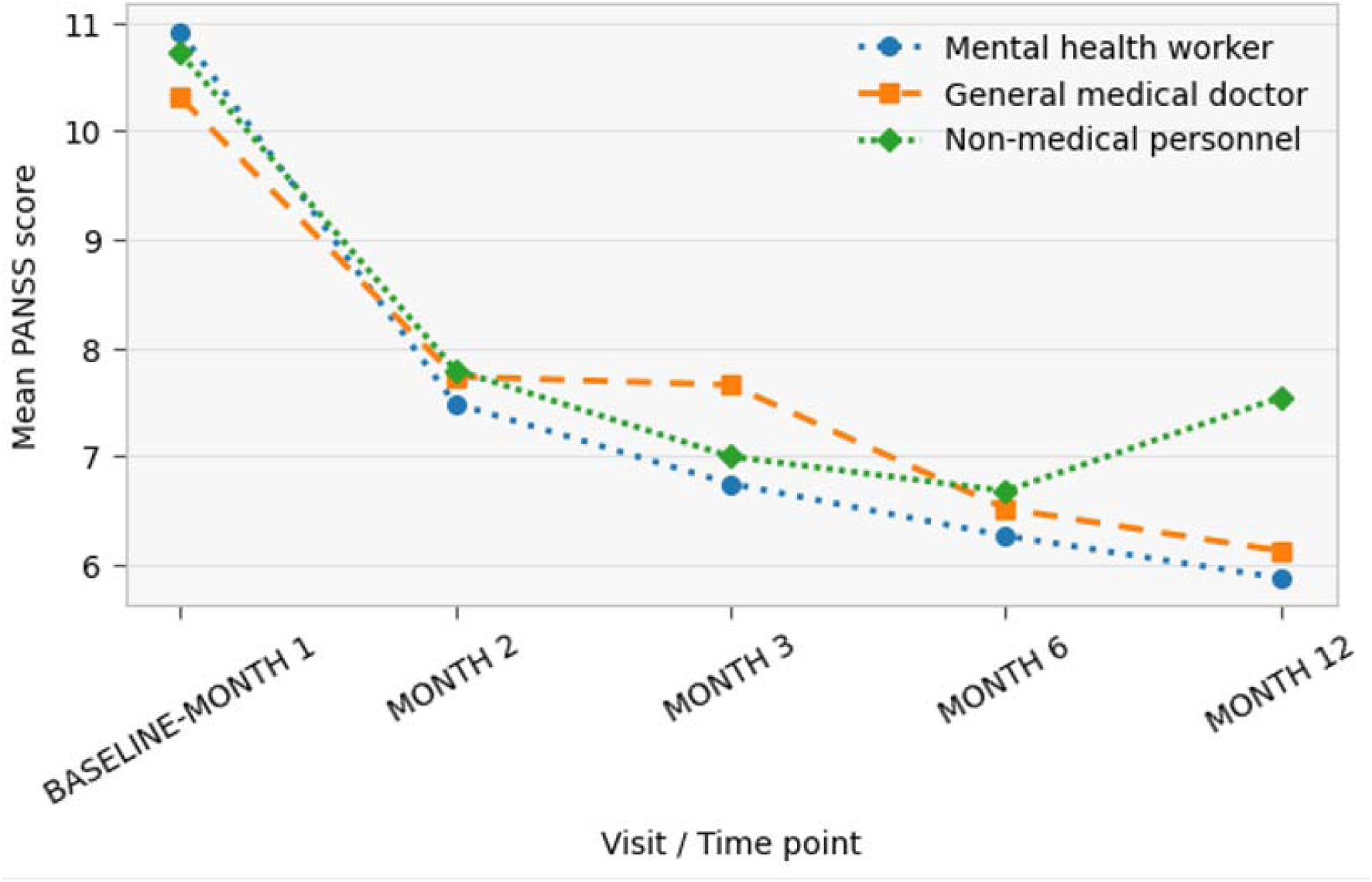
Changes in PANSS Scores Over Time by Initial Care Provider.

**Figure 4:**
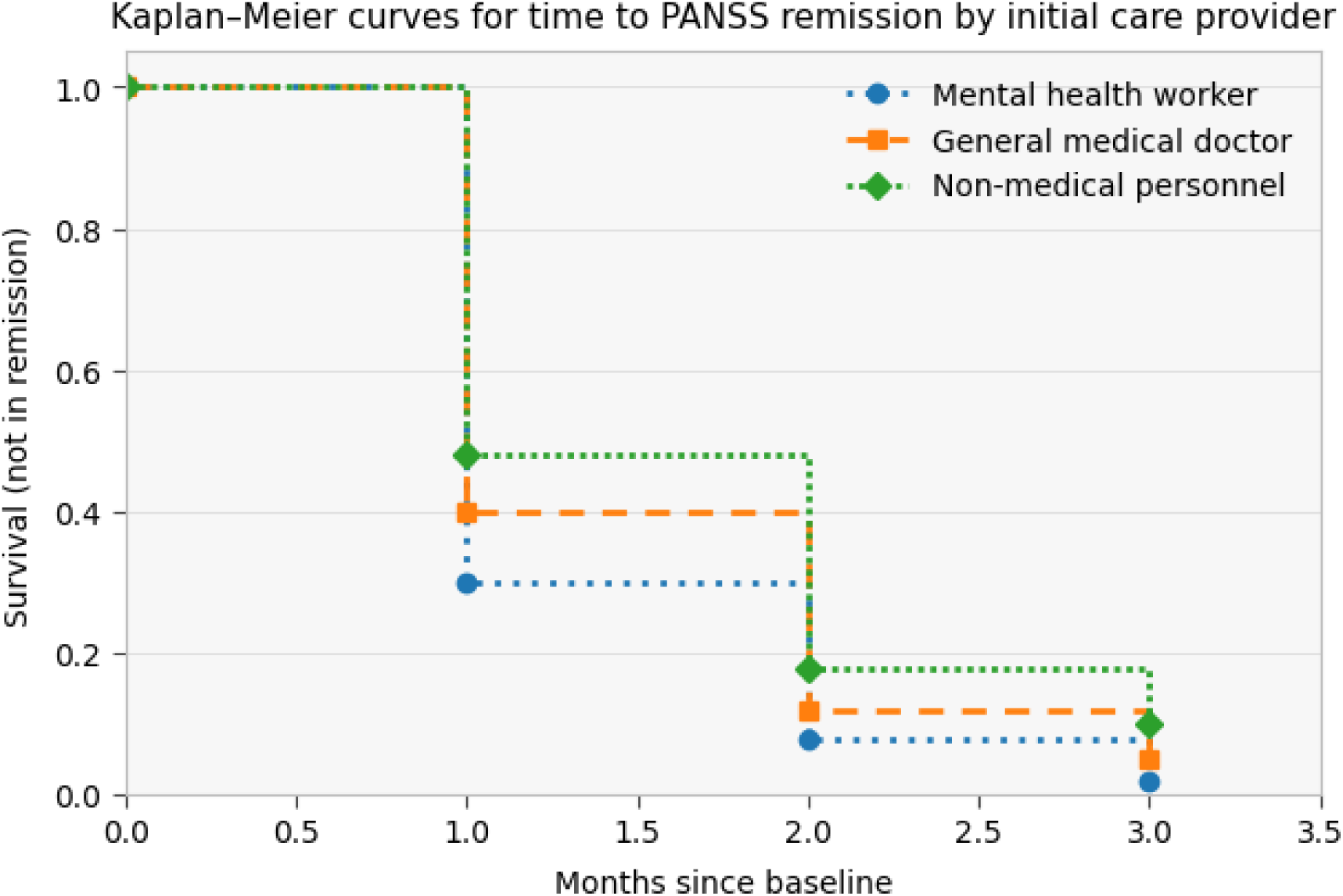
Time to symptom improvement by initial provider.

Over the follow-up period, the cumulative probability of remaining non-remitted declined in all groups. Participants initially seen by mental health workers exhibited the lowest survival probabilities (i.e., the highest cumulative incidence of remission) at each follow-up time point, followed by those initially seen by general medical doctors, whereas participants initially seen by non-medical personnel had the highest probability of remaining non-remitted. Divergence between the curves was observed early in follow-up and persisted across subsequent time points, although the probability of non-remission was low in all groups by the end of follow-up

### Adjusted Cox proportional hazards model for predictors of time to symptom remission

In the adjusted Cox proportional hazards model, the initial care provider was associated with time to PANSS remission. Compared with participants initially seen by non-medical personnel, those initially managed by mental health workers had a higher rate of remission (adjusted HR = 1.48; 95% CI: 1.05–2.10; p = 0.026). Participants initially seen by general medical doctors did not differ significantly from those seen by non-medical personnel (adjusted HR = 1.12; 95% CI: 0.78–1.61; p = 0.540). Baseline symptom severity was strongly associated with remission; higher baseline PANSS scores were associated with a lower rate of remission (adjusted HR = 0.91 per point increase; 95% CI: 0.87–0.95; p < 0.001). Increasing age was also associated with a slower rate of remission (adjusted HR = 0.94 per year; 95% CI: 0.91–0.97; p = 0.023). Sex and body mass index were not significantly associated with time to PANSS remission

**Table 3:** Predictors to symptom remission.

| Covariate | Adjusted HR | 95% CI | p-value |
| --- | --- | --- | --- |
| **Initial care provider** |  |  |  |
| Mental health worker | **1.48** | **1.05 – 2.10** | **0.026** |
| General medical doctor | 1.12 | 0.78 – 1.61 | 0.540 |
| Non-medical personnel | Reference |  |  |
| Baseline PANSS score (per point) | 0.91 | 0.87 – 0.95 | <0.001 |
| Age (per year) | 0.94 | 0.91 – 0.97 | 0.023 |
| Male sex | 1.01 | 0.58 – 1.76 | 0.981 |
| BMI (per kg/m<sup>2</sup>) | 1.00 | 0.98 – 1.02 | 0.921 |

### Quality of life

Mean quality-of-life (QOL) scores increased over time across all three initial care provider groups. At baseline, mean QOL scores were similar across groups, ranging from approximately 5.6 to 5.7. By Month 2, QOL scores had increased across all groups, with participants initially seen by non-medical personnel showing slightly higher mean scores than those initially seen by mental health workers or general medical doctors. Between Month 2 and Month 3, a marked increase in mean QOL was observed among participants initially seen by mental health workers, reaching the highest mean QOL score at Month 3. Continued improvement was observed through Month 6 across all groups, with mean QOL scores converging at approximately 7.2–7.6. By Month 12, mean QOL scores remained highest among participants initially seen by mental health workers, followed by those initially seen by general medical doctors, while participants initially seen by non-medical personnel had slightly lower mean scores

**Figure 5:**
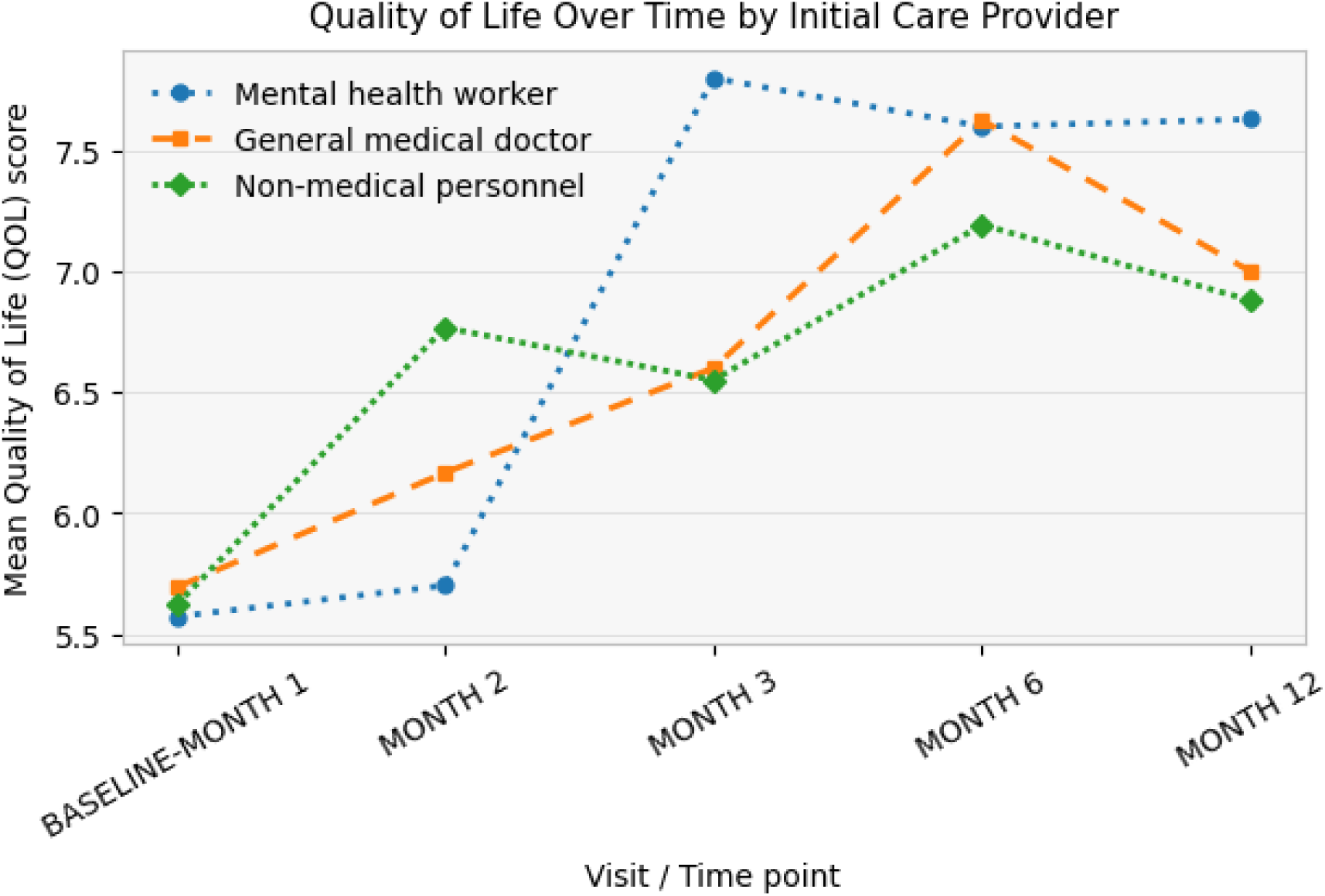
Quality of life changes over time by initial care provider.

#### Time-to-Event Analysis

Kaplan–Meier curves were used to describe time to achieving good quality of life (defined as QOL ≥ 7) by initial care provider. Across follow-up, the probability of not yet achieving good QOL declined in all groups. Participants initially seen by mental health workers consistently showed the lowest probability of remaining below the QOL threshold, followed by those initially seen by general medical doctors, while participants initially seen by non-medical personnel had the highest probability of remaining below the threshold.

**Figure 6:**
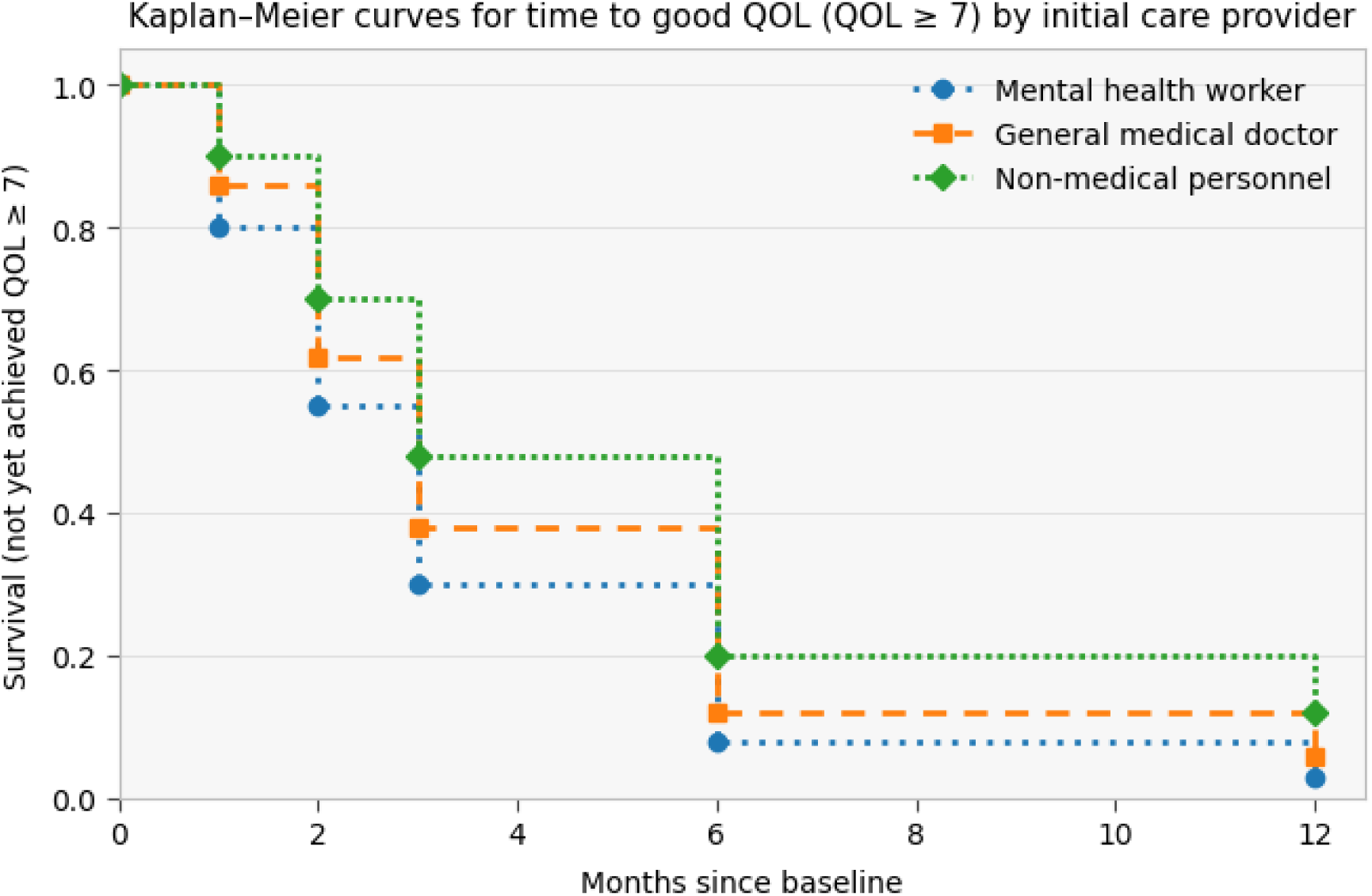
KM graph for time to good quality of life by initial care provider.

#### Cox Proportional Hazards Model

A Cox regression model was fitted with age, sex, BMI, PANSS, as covariates. Hazard ratios (HR) greater than 1 indicate faster achievement of satisfaction; HR less than 1 indicate slower time. Compared with participants initially seen by non-medical personnel, those initially managed by mental health workers had a higher rate of achieving good QOL (adjusted HR = 1.36; 95% CI: 1.02–1.82; p = 0.033). Participants initially seen by general medical doctors did not differ significantly from those initially seen by non-medical personnel (adjusted HR = 1.10; 95% CI: 0.82–1.48; p = 0.540). Higher baseline QOL scores were associated with a faster transition to good QOL (adjusted HR = 1.22 per point increase; 95% CI: 1.10–1.35; p < 0.001), while higher baseline PANSS scores were associated with a slower transition (adjusted HR = 0.94 per point increase; 95% CI: 0.90–0.98; p = 0.004). Age, sex, and body mass index were not significantly associated with time to achieving good QOL.

| Covariate | Adjusted HR | 95% CI | p-value |
| --- | --- | --- | --- |
| Initial care provider |  |  |  |
| Mental health worker | 1.36 | 1.02 - 1.82 | 0.033 |
| General medical doctor | 1.1 | 0.82 - 1.48 | 0.54 |
| Non-medical personnel | Reference |  |  |
| Baseline QOL score (per point) | 1.22 | 1.10 - 1.35 | <0.001 |
| Baseline PANSS score (per point) | 0.94 | 0.90 - 0.98 | 0.004 |
| Age (per year) | 0.99 | 0.96 - 1.02 | 0.52 |
| Male sex (vs female) | 0.97 | 0.72 - 1.31 | 0.85 |
| BMI (per kg/m<sup>2</sup>) | 1 | 0.98 - 1.02 | 0.91 |

## Discussion

This study provides the first empirical examination of referral pathways and their impact on patient outcomes in individuals with a first episode of psychosis to Uganda’s first Early Intervention in Psychosis Service, the Specialized Treatment Early in Psychosis Clinic at Makerere University Hospital (STEP_MaKH). Three key findings emerged. First, help-seeking trajectories were highly variable and often began outside the formal health system, particularly with traditional and religious healers. Second, fewer than one-third of eligible patients accepted referral to specialised early psychosis care, with referral site, initial care provider, and delays from first contact strongly predicting referral completion. Third, although clinical outcomes improved substantially over 12 months for all participants, outcomes were better when initial contact was with a mental health worker. These findings have important implications for scaling early psychosis care in low-resource settings.

### Pathways to care

Pathways were highly variable and frequently began outside formal psychiatric care. Nearly half of the participants first sought help from traditional or religious healers, while others entered care through general psychiatric services, general hospitals, or (less commonly) the police or social services. This pattern is consistent with evidence across sub-Saharan Africa showing that explanatory models of illness, stigma, access barriers, and health-system fragmentation produce non-linear, multi-contact trajectories before specialist care is reached (18, 20, 21, 26). Importantly, the pathway diagrams also illustrate “cycling” across provider types, suggesting that delays are not simply due to late first help-seeking, but to repeated transitions between providers before effective linkage to specialised services.

### Referral Patterns

There was substantial attrition before STEP_MaKH. Fewer than one-third of eligible patients completed referral, and predictors were predominantly system- and pathway-level features rather than basic demographics. Referral site was the strongest determinant, with referral completion being substantially higher at Mulago than at Butabika. The unacceptably long delay in returning of mental health services to Mulago Hospital after renovation may be the reason why there was greater referral. Initial contact at a general hospital was also associated with reduced probability of enrolling at STEP_MaKH, plausibly reflecting competing care pathways, limited structured referral mechanisms, or weak continuity across facility types. Beyond the site, longer delays from first contact reduced the likelihood of reaching STEP_MaKH, indicating that time itself functions as an attrition mechanism (27, 36). Possible explanations include families becoming demoralised, resources being depleted, symptoms becoming chronic, or patients stabilising sufficiently in non-specialist settings that onward referral is deprioritised (37–39). In the adjusted model, these results suggest that scaling EIPS in Uganda could be improved by strengthening referral systems and the reliability of linkages, particularly in high-volume psychiatric settings where most patients first present.

### Pathways and Clinical Outcomes

Patients initially seen by mental health providers on average had more severe illness severity. This is in keeping with previous literature from this setting, highlighting how patients with more severe psychosis are sent to psychiatric services (40). Once patients entered STEP_MaKH, outcomes improved rapidly, with initial provider type showing measurable associations with speed of improvement. Irrespective of initial provider type, symptom severity declined rapidly, with 60% achieving PANSS remission (scores < 14) by Month 1 and very low non-remission by Months 2–3. This is similar to high-income literature highlighting the potential of specialised early psychosis care even in low-resource contexts when a functional pathway into care exists (14, 26). However, initial contact with a mental health worker was associated with a shorter time to PANSS remission than contact with non-medical personnel, and mental health worker contact similarly predicted earlier attainment of good QOL. The general medical doctor pathway did not differ significantly from non-medical personnel in either model, suggesting that the benefit is not simply “any medical contact,” but may reflect earlier recognition of psychosis by trained mental health providers. Baseline severity strongly predicted slower remission, indicating that early-stage severity remains a key driver of trajectory even after specialised care begins (41, 42). Older age predicted slower remission, which may reflect longer untreated illness, more entrenched functional impairment, differential family responses, or unmeasured comorbidity (43, 44). For quality of life, higher baseline QOL accelerated attainment of good QOL, while higher baseline PANSS slowed it, reinforcing the clinical logic that symptom burden and functional recovery remain tightly coupled (45, 46).

### Implications for Early Intervention in Low-Resource Settings

Improving early psychosis outcomes in Uganda and across sub-Saharan Africa requires integrating clinical evidence, system design, and practical implementation. STEP_MaKH demonstrates that specialized early intervention services produce tangible benefits in symptom resolution and quality of life, even in resource-constrained settings. Their effectiveness depends critically on the pathways that lead to them. Differences in referral completion rates between Mulago and Butabika hospitals reveal high-impact opportunities for system optimization: standardized referral protocols, rapid family orientation at point of referral, warm handoffs with scheduled appointments, and bidirectional feedback loops that enable referring providers to track outcomes and sustain referral behaviour. These mechanisms shorten the time between first help-seeking and specialist linkage. This system optimization has been done in existing early intervention services and networks including EPINET, Australian Early Psychosis Collaborative Consortium (AEPCC) and Early intervention services in the UK (35, 47, 48). Similar services must be adapted to low resource settings in Africa. Since traditional and religious healers often serve as the first point of contact, they must be engaged as partners rather than obstacles (22, 23). This may be through co-developed referral agreements, culturally sensitive psychoeducation, and shared communication channels that facilitate timely transitions to formal care. For sustainable scale-up, early intervention programs must be embedded within national mental health strategies and existing health system structures, not developed in isolation. However, coordinating nodes for various services are required. Other practical implementation steps include: adopting digital referral-tracking systems to identify bottlenecks in real time, building capacity among frontline mental health workers to accelerate both linkage and clinical recovery, aligning program placement with institutional readiness and community demand, and establishing measurement-based care platforms that support ongoing quality improvement (49–51).

### Future Directions

Three research priorities will advance early psychosis care in sub-Saharan Africa. First, establish standardized measurement across diverse contexts. Multi-site cohorts spanning varied health systems and cultural settings are needed to move beyond single-site findings. Standardized tools like the Nottingham Onset Schedule (NOS-DUP) should measure duration of untreated psychosis, enabling clearer interpretation of treatment delays and meaningful comparison with global benchmarks (52). Second, understand the human factors shaping care pathways. Qualitative research with patients, caregivers, traditional healers, and frontline clinicians should examine how cultural beliefs, stigma, economic barriers, and trust in different care providers influence help-seeking decisions and pathway navigation. This work must identify not just barriers but also existing strengths in community support systems that can be leveraged. Third, test implementation strategies through pragmatic trials. Rather than demonstrating efficacy alone, studies should rigorously evaluate scalable interventions including digital referral tracking, community-based psychoeducation co-designed with traditional healers, task-shifting models for frontline workers, and measurement-based care protocols. Their effectiveness should be compared across different facility types and referral networks. These trials should measure not only clinical outcomes but also implementation metrics: referral completion rates, time to first specialist contact, cost per patient linked, and sustainability beyond external funding. In all these phases, the inclusion of people with lived experience will ensure the interventions are acceptable and useful (53). Together, this research agenda will generate the context-specific evidence needed to reduce treatment delays and expand access at scale.

### Strengths and Limitations

A major strength of this study is its use of structured instruments, such as the WHO Encounter Form, PANSS-6, and WHO-QOL, to capture both pathway dynamics and longitudinal clinical outcomes. By focusing on Uganda’s first EIPS, the study provides novel data from a context where such services are still in their early stages of development. However, several limitations warrant caution. The sample size for patients enrolled at STEP_MaKH was modest, limiting statistical power for some outcomes. Pathway reconstruction relied on retrospective reporting, raising the possibility of recall bias. The study did not directly measure DUP using validated instruments such as the NOS-DUP; however, DUP is likely a key mediator of the observed associations between pathways and outcomes. Finally, findings reflect patterns in a specific urban referral context and may not generalise to rural settings or other regions of Uganda.

## Conclusion

In Uganda, as in much of sub-Saharan Africa, pathways to care in patients with psychotic disorders vary and are culturally mediated. However, empirical evidence linking these pathways to health-system attrition and downstream outcomes has been limited. This study provided some answers by examining referral completion into STEP_MaKH alongside 12-month clinical trajectories after enrolment. Our findings show that specialised early intervention can produce rapid clinical gains, but that its population impact depends on whether patients can reliably reach it early enough for those gains to matter. The dominant bottleneck is referral completion, which was low and was shaped primarily by the referring site and by cumulative delays from first contact. Once patients entered specialised care, such as STEP_MaKH, outcomes improved rapidly. However, earlier contact with mental health workers predicted faster remission and earlier quality-of-life gains, suggesting that pathway “quality” and not just pathway “length,” influences outcomes. Importantly, these results reposition scale-up as an implementation problem requiring standardised protocols at key entry points, active linkage and navigation for families, and targeted capacity building for frontline mental health workers. These interventions may reduce attrition, shorten delays, and accelerate recovery in real-world African health systems.

## Data Availability

All data produced in the present study are available upon reasonable request to the authors.

## Ethical approvals

Ethical approval was obtained from the School of Medicine Research and Ethics Committee of Makerere University College of Health Sciences (Mak-SOMREC-2024-869). Institutional approval was also obtained from Butabika National Referral Mental Hospital. All participants provided informed consent.

## Author Contribution

EKM drafted the manuscript. EKM, RK, MK,MS and NN contributed to the conceptualisation of the study design. All co-authors have revised the manuscript critically for important intellectual content.

## Acknowledgements

We are grateful the Brain Health Study that supported this grant. Special appreciation to the participants for providing their information.

